# Systemic inflammation during fasting and postprandial states: a comprehensive study of key determinants in a deeply characterized cohort of young adults

**DOI:** 10.1101/2024.08.30.24312659

**Authors:** Parvaneh Ebrahimi, David Horner, David Burgner, Nicklas Brustad, Tingting Wang, Mina Ali, Liang Chen, Ann-Marie M Schoos, Klaus Bønnelykke, Jakob Stokholm, Evrim Acar, Nilo Vahman, Bo Chawes, Morten A. Rasmussen

## Abstract

Systemic inflammation contributes to the pathogenesis of many noncommunicable diseases. Additionally, postprandial inflammation can exacerbate systemic inflammation. These emphasize the need to examine inflammation in both fasting and postprandial states, to identify modifiable factors to alleviate inflammation. This study investigated a comprehensive list of factors spanning from foetal stage to young-adulthood against inflammation levels at fasting (chronic inflammation) and postprandial states (meal-induced transient inflammation). A meal challenge was undertaken in 18-year-olds *(n* = 298), and inflammation was assessed using the robust GlycA biomarker. Associations between inflammation and various factors were observed, some of which were sex-specific; e.g. the associations of alcohol consumption and smoking with inflammation were significantly stronger in females. Moreover, novel associations from gestation and early life (e.g. pregnancy smoking) were identified. Our findings highlight factors that should inform dietary and lifestyle interventions for reducing systemic inflammation and highlight the importance of considering inflammation in precision nutrition practices.

## Introduction

Systemic inflammation is a key underlying mechanism of many chronic non-communicable diseases, including cardiovascular disease (CVD), type 2 diabetes (T2D), allergic diseases, and asthma [1-4]. Recognised drivers of local and systemic inflammation include host genetics [5], and environmental exposures including dietary intake [6]. The relationship between diet and inflammation extends beyond the effects of prolonged exposure; notably, the consumption of a single meal can also lead to an acute increase in inflammatory markers, a phenomenon referred to as ‘postprandial inflammation’ [7]. The repetitive postprandial inflammatory state, characterized by elevated triglycerides and glucose, promotes oxidative stress and inflammatory processes, further exacerbating systemic inflammation [8].

The composition of a given meal affects postprandial response, with high-fat/high-sugar meals triggering a more intense postprandial (inflammatory) response [9], which is the basis for meal-challenges such as Oral Glucose Protein Lipid Tolerance Test (OGPLTT). A high-fat diet is intricately linked to increased CVD risk and insulin resistance [10]. On the other hand, inflammation is associated with both CVD risk and insulin resistance, suggesting its potential role as a mediator in the effects of a high-fat diet [11, 12].

During fasting, metabolism operates at a steady state, whereas the postprandial phase involves significant metabolic changes as the body processes nutrients, resulting in various physiological responses, including postprandial inflammation. In otherwise healthy individuals, systemic inflammation in the fasting state reflects mostly chronic inflammation, whereas systemic inflammation after the consumption of a meal reflects acute-phase postprandial inflammation [13]. Although chronic inflammation is studied to a larger extent, postprandial inflammation and its relationship with disease remains under-explored. However, there is value in investigating postprandial phase and inflammation more in depth, as many individuals spend most of their day in a postprandial state. Besides, inter-individual variations may not be fully evident in the more metabolically-steady fasting state, and postprandial inflammation can show greater variability than fasting [14], as the host metabolism is challenged by the meal. The fact that identical meals can elicit varied postprandial responses among individuals highlights the potential of taking a precision nutrition approach towards identifying the determinants and predictors of systemic inflammation at the postprandial state as well.

A relatively newly described inflammatory biomarker is glycoprotein acetylation (GlycA), that provides a composite measure of systemic inflammation [15]. GlycA has proved to be of high clinical relevance, and has been linked to metabolic disorders, including diabetes and obesity [16-20]. GlycA is robust and has low intra-individual and intra-assay variability. Though, the main virtue of GlycA that makes it a good candidate for measuring inflammation at metabolic states is the fact that it is consistently responsive to dietary intake [21, 22], which is not the case for most of the other more conventional inflammatory markers, including C-reactive protein (CRP) [8, 23]. Therefore, GlycA is a promising biomarker for assessing postprandial inflammatory response.

In this study, we investigate the association of a comprehensive and diverse range of (risk) factors with: 1) chronic inflammation during fasting state, and 2) postprandial inflammation after a standardized OGPLTT meal-challenge, in a prospective mother-child cohort of generally healthy young adults [24, 25]. The investigated factors span from gestation into adolescence, including prenatal and early-life exposures, genetics, lifestyle, environmental factors, blood pressure, clinical laboratory tests, etc., and we use GlycA to assess the intensity of inflammation. Furthermore, we investigate insulin resistance, as a key component of metabolic health, and show how inflammation both mediates and moderates the effect of well-known anthropometric risk factors on insulin resistance. Our results show differences between sexes and highlight some of the factors that should be considered in providing dietary and lifestyle interventions, to help ameliorate chronic and postprandial inflammation.

## Results

### The study design

An OGPLTT was undertaken by 298 generally healthy 18-year-olds (154 females; 144 males), from the COPSAC2000 cohort that includes 410 individuals in total. Blood samples were collected and analysed by NMR to measure fasting and postprandial inflammation (0– 240 min) (Methods). Figure 1 shows the experimental design, and the cohort characteristics of the participants are presented in Supplemental Table 1. Among the cohort population, except for Homeostatic Model Assessment for Insulin Resistance (HOMA-IR) which was higher in non-participants, other characteristics were not significantly different between the participants versus non-participants.

**Fig 1.**
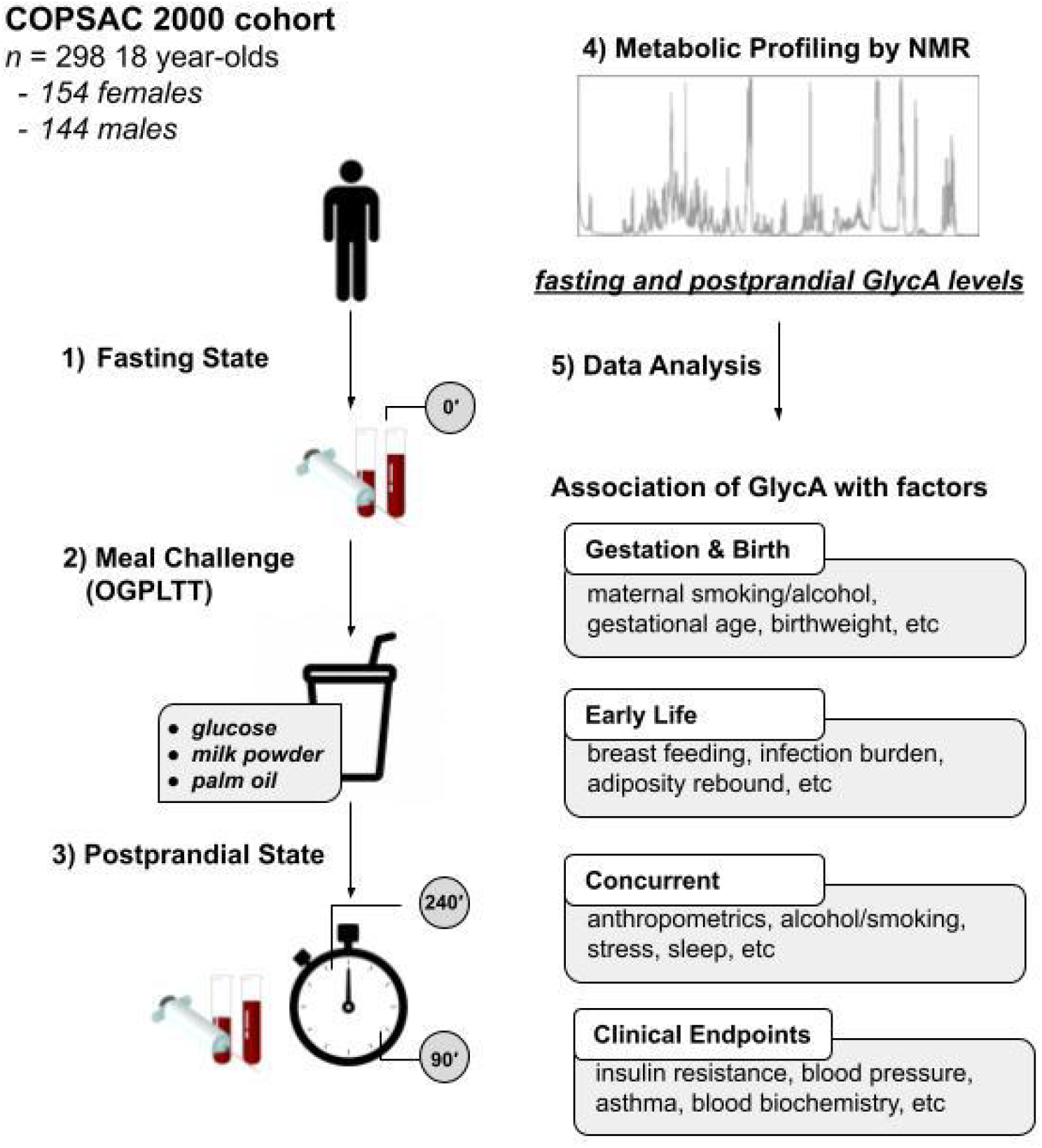
Conceptual overview of the study.

### Characteristics of fasting and postprandial inflammation

Fig. 2 shows dynamics and changes of GlycA, including fasting (T = 0 min) and the seven postprandial time points. For both sexes a postprandial increase in GlycA was observed, measured as the difference between fasting and the peak postprandial GlycA concentration. However, 11% (*n* = 17) of females and 8% (*n* = 11) of males showed no increase. The median fasting GlycA concentration was 5.6% higher in females (*P* = 3.1*e*^-3^), and females on average had higher GlycA levels at all the postprandial time points. However, males had on average a more intensified postprandial increase in GlycA levels relative to fasting, with the median increase in males being 7.5% versus 5.5% (*P* = 0.01) in females. The median GlycA concentration peaked at 60 min in females, and at 90 min in males. Looking at individual observations, in both sexes, postprandial inflammation peaked at 60 min for most subjects, followed by 90 min in males and 30 min in females.

**Fig 2.**
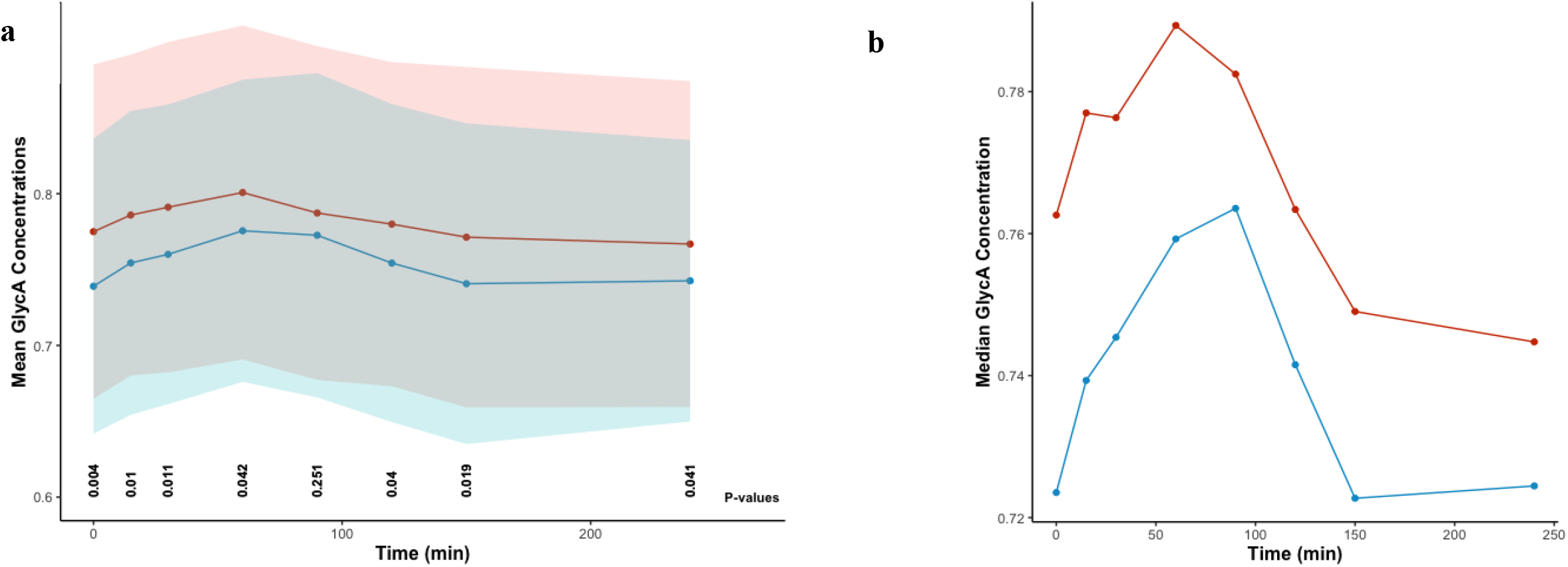
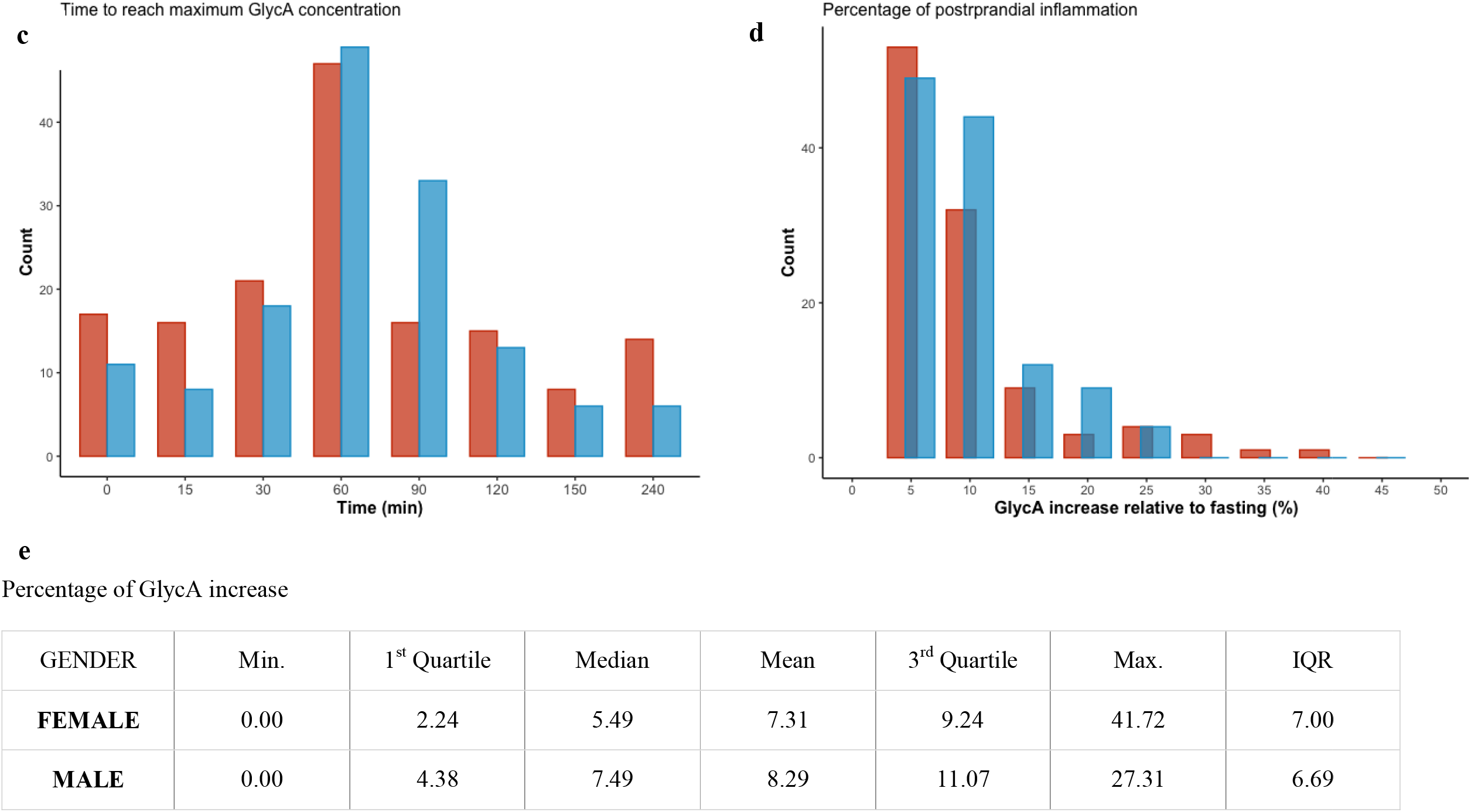
Sex-stratified postprandial dynamics of GlycA. a) mean GlycA time-profiles with standard deviation ribbons, and p-values for the differences between sexes; b) median GlycA levels at fasting and postprandial time-points; c) histogram of time to peak GlycA concentrations; d) percentage of increase in GlycA concentrations, calculated as the difference between the maximum postprandial GlycA concentration and the fasting level, divided by the fasting GlycA level; e) descriptive statistics for the percentage of GlycA increase. ‘Red’ visualizations denote females, while ‘Blue’ visualizations represent males.

### Predictors of fasting and postprandial inflammation

GlycA levels at fasting and two postprandial time-points were selected for further analysis. From the postprandial phase 90 min and 240 min were chosen (early and late postprandial stages respectively, chosen based on the median GlycA profiles), to incorporate the diet-induced inflammatory response as well as the clearance phase. As excess adiposity is known to be a key determinant of obesity-related inflammation, linear models were fitted for fasting levels and the incremental area under the curve (iAUC) of GlycA versus body fat percentage (Supplemental Fig. 1), to investigate this relation in our cohort. Fasting GlycA levels were significantly associated with fat percentage in both sexes, however with stronger inference in females (*R*^2^ = 0.25 ; *P* < 10^−10^) compared to males (*R* ^2^ = 0.12 ; *P* < 10^−4^). Postprandial GlycA levels and adiposity showed weaker associations with only marginal significance in males at 240 min, but not significant in females.

Fasting and postprandial GlycA levels were modelled against a range of factors categorized as pregnancy (*k* = 5 factors), birth (*k* = 4), early life (*k* = 10), current (*k* = 10), and clinical endpoints (*k* = 11), using (multiple) linear regression. The aim was to assess the association of these risk factors with the individuals fasting and postprandial inflammation. Depending on the aetiology of the risk factor, univariable or multivariable linear models were fitted, and for anthropometrics and genetic risk, backward selection was used to identify the most significant predictors.

For certain factors, stratification by sex was deemed necessary due to biological influences, or stratification revealed sex-specific differences not apparent in non-stratified models. These factors included adiposity rebound, Tanner score, anthropometrics, genetics, current smoking, alcohol consumption, accelerometery, HOMA-IR, and blood pressure. The list of the risk factors, the rationale behind the stratification of some of them, as well as the complete results of the models are presented in Supplemental Table 2.

The key associations during fasting and postprandial states are outlined below, initially focusing on non-stratified factors (models), followed by sex-stratified models.

### a) Fasting-state inflammation

Fig. 3a summarizes the linear models of the predictors of fasting GlycA levels. Effect sizes in the models were estimated using (adjusted) coefficient of determination (*R*^2^) for univariable and multiple linear models. In the models that included adjustment variables, an Analysis of Variance (ANOVA) was subsequently performed, to calculate ‘eta squared’ (*η*^2^ : sum of squares of the predictor/total sum of squares), as an estimate of the effect size.

**Fig 3.**
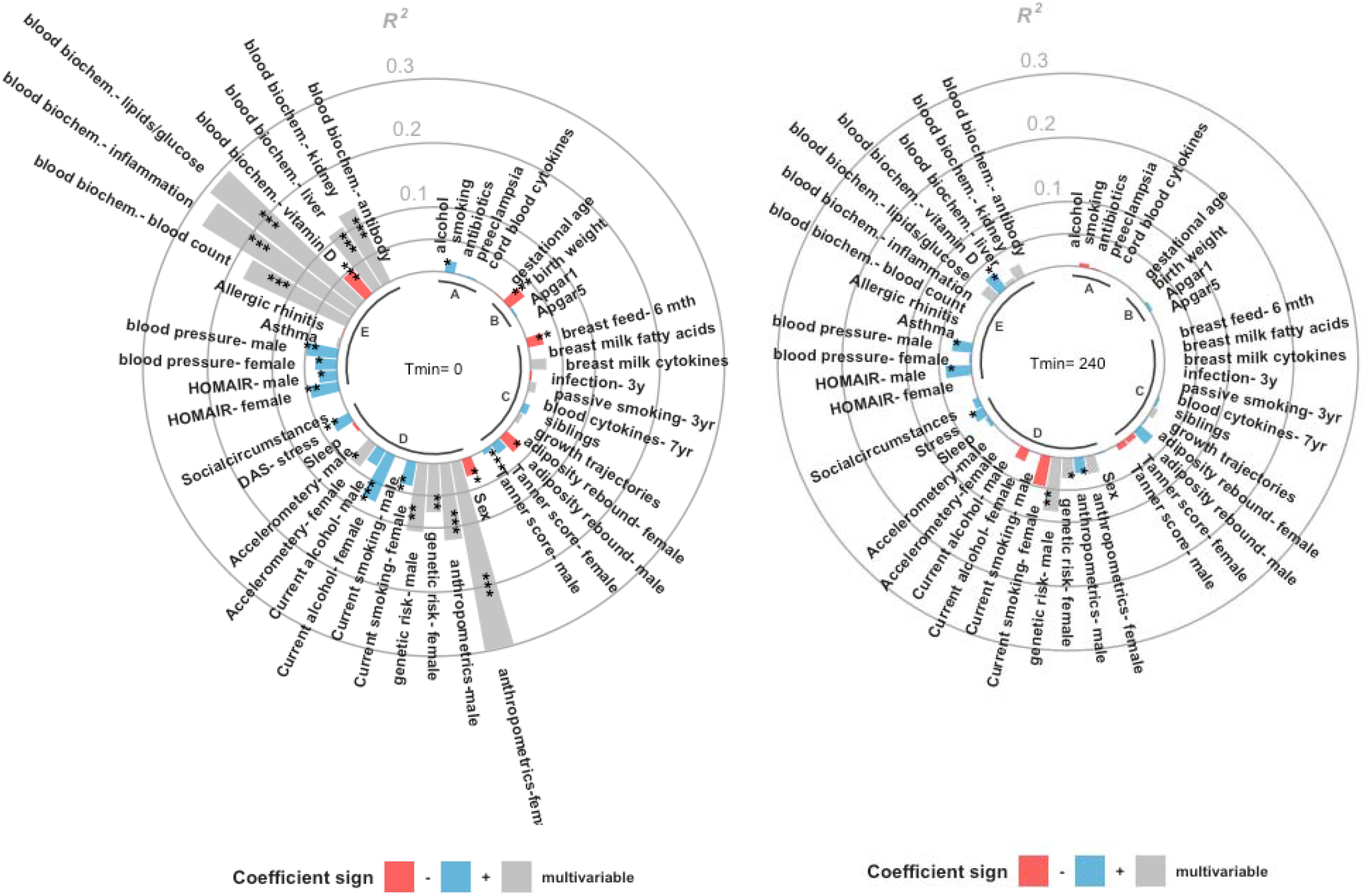
Determinants of fasting and postprandial GlycA. a) fasting state, b) postprandial state (GlycA iAUC_240min_). The bars show the effect sizes ‘Blue’ bars represent positive regression coefficients as acquired from univariable linear regression, while ‘Red’ bars represent negative regression coefficients; these correspond to positive and negative associations between the risk factor and GlycA levels, respectively. For risk factors with multivariable models, the bars are colored ‘Gray’, with no immediate direction of association obtained. The labeled components in the figure correspond to: A) gestation, B) birth, C) early-life, D) current, and E) clinical endpoints.

Current factors and clinical laboratory tests included the strongest associations with fasting GlycA. Clinical blood biochemistry measures from fasting state showed the highest association, with lipids/glucose (cholesterol, triglycerides, glycated haemoglobin (HbA1c), and glucose), inflammation (hs-CRP and erythrocyte sedimentation rate), and blood cell count categories being the strongest (*R*^2^ = 0.29, 0.27, *and* 0.18 ; *P* < 10^−10^). 25-hydroxy vitamin D measures of blood showed negative association (*R*^2^ = 0.05 ; *P* < 10^−3^). The list of the investigated blood tests is included in Supplemental Table 3.

Factors from gestation and early life were also associated with fasting GlycA. Pregnancy smoking, adjusted for current smoking and social circumstances, was associated with increased GlycA levels (*η* ^2^ = 0.02 ; *P* = 0.02), while birth weight adjusted for current BMI (η^2^ = 0.03 ; *P* < 10^−3^), and being breastfed for at least the first 6 months of life (*R*^2^ = 0.02 ; *P* = 6.7*e*^-3^) showed negative association with fasting GlycA. Furthermore, age at adiposity rebound showed negative association with GlycA levels in males, remaining marginally significant after adjusting for current BMI (*η*^2^ = 0.03 ; *P* = 0.04).

Some of the sex-stratified models also showed associations with fasting GlycA. Overall, the levels of fasting GlycA were higher in females (*R*^2^ = 0.03 ; *P* = 3.9*e*^-3^). In females, anthropometric measures of body fat percentage, waist/hip, muscle/fat, and waist/height (identified by backward elimination) showed high positive association with fasting GlycA (*R*^2^ = 0.30; *P* < 10^−10^). However, in males, the effect of anthropometrics was captured solely by BMI (identified by backward elimination) as a weaker predictor (*R*^2^ = 0.12; *P* < 10^−4^). To assess the influence of host genetics, the associations between fasting GlycA and 15 polygenic risk scores (PRS) for traits that are linked to inflammation or metabolism (including CVD, T2D, allergy, etc; Supplemental Table 4) were investigated. Using a multivariable model with backward elimination, seven PRS were selected in males (‘basal metabolic rate’ and ‘serum gamma-glutamyl transferase measurement’ the most significant) and five in females (‘HbA1c measurement’ the most significant). These selected PRS associated positively with fasting GlycA, explaining 11% and 8% of the variance respectively in males and females (P < 0.01). Current life-style factors showed sex-specific associations. Heavier alcohol consumption (≥ 10 units/week) in females was associated with significant increase in fasting GlycA levels (*R*^2^ = 0.08 ; *P* < 10^−3^), though only marginally significant in males (*R*^2^ = 0.03 ; *P* = 0.05); this association remained significant even after correcting for body fat percentage to account for the potential role of adiposity. Moreover, smoking in females was positively associated with fasting GlycA levels (*η*^2^ = 0.04 ; *P* = 5.3*e*^-3^), but was not significant in males.

Physical activity, as estimated by wrist-worn accelerometer, showed significant association in females (*R*^2^ = 0.04 ; *P* = 0.03). The model included light, moderate, and vigorous activity measures, with vigorous activity showing the strongest inverse association with GlycA.

Higher diastolic blood pressure was associated with higher fasting GlycA in both sexes, with a more pronounced effect in males accounting for 4.8% (*P* = 8.2*e*^-3^) of the variance compared to 3.4% (*P* = 0.02) in females. Systolic blood pressure (not shown in Fig. 3) showed a similar association in females (*R*^2^ = 0.03 ; *P* = 0.03), though not significant in males. Moreover, HOMA-IR had positive association with GlycA, in both females (*η*^2^ = 0.04 ; *P* = 3.0e^-3^), and males (*η*^2^ = 0.03 ; *P* = 0.04).

### b) Postprandial inflammation

Fig. 3b summarizes the linear models of the predictors of postprandial GlycA levels at 240 min as measured by incremental area under the curve, taking the baseline levels into account. The results for 90 min are shown in Supplemental Fig. 2.

Most of the factors showing associations with postprandial GlycA at 240 min were sex-specific. Genetic risk in males showed the strongest association with GlycA iAUC_240min_, where PRS for ‘T2D’ and ‘basal metabolic rate’ explained 8% of the variance (*P* = 2.0e^-3^). In females, this association was weaker, with PRS for ‘insulin resistance’ and ‘hypothyroidism’ explaining 3% of the variance (*P* = 0.04). HOMA-IR in males showed positive association (*η*^2^ = 0.04 ; *P* = 0.02), though not significant in females. Furthermore, stress was positively associated with GlycA iAUC_240min_ (*R*^2^ = 0.02 ; *P* = 0.03), whereas diastolic blood pressure was positively associated only in males (*R*^2^ = 0.03 ; *P* = 0.03). Anthropometric measures were also significant only in males, where waist-to-height ratio (backward elimination) showed positive association (*R*^2^ = 0.03 ; *P* = 0.03). 25-hydroxy vitamin D measures of blood with positive association (*R*^2^ = 0.03 ; *P* = 2.1*e*^-3^) was the only blood biochemistry measure that was significant.

Only a few of the investigated risk factors were significantly associated with postprandial GlycA at 90 min. 25-hydroxy vitamin D measure was positively associated with GlycA iAUC_90min_ (*R*^2^ = 0.04 ; *P* < 10^−3^). Psychological stress was also positively associated with GlycA iAUC_90min_ (*R*^2^ = 0.02 ; *P* = 0.04). From the sex-stratified models, genetic risk was only significant in males where PRS for ‘T2D’ and ‘basal metabolic rate’ (selected by backward elimination) explained 9.2% of the variance (*P* = 1.1e^-3^).

### Prediction of GlycA levels using fasting and postprandial metabolome

Linear models were used to explore the relationship between fasting concentrations of 12 groups of metabolites and fasting GlycA levels, aiming to understand how the fasting metabolome corresponds to inflammation (Fig. 4). The strongest association was for ‘*Other lipids category*’, which included sphingomyelins, phosphoglycerides, cholines, etc. (grouping of metabolites defined based on Nightingale analytical platform definitions; see Supplemental Table 5); this category explained 51% of the variance in fasting GlycA. The second strongest groups were fatty acids, triglycerides, and phospholipids corresponding to 32-46% of the variance, whereas glycolysis related metabolites explained only 18%.

**Fig 4.**
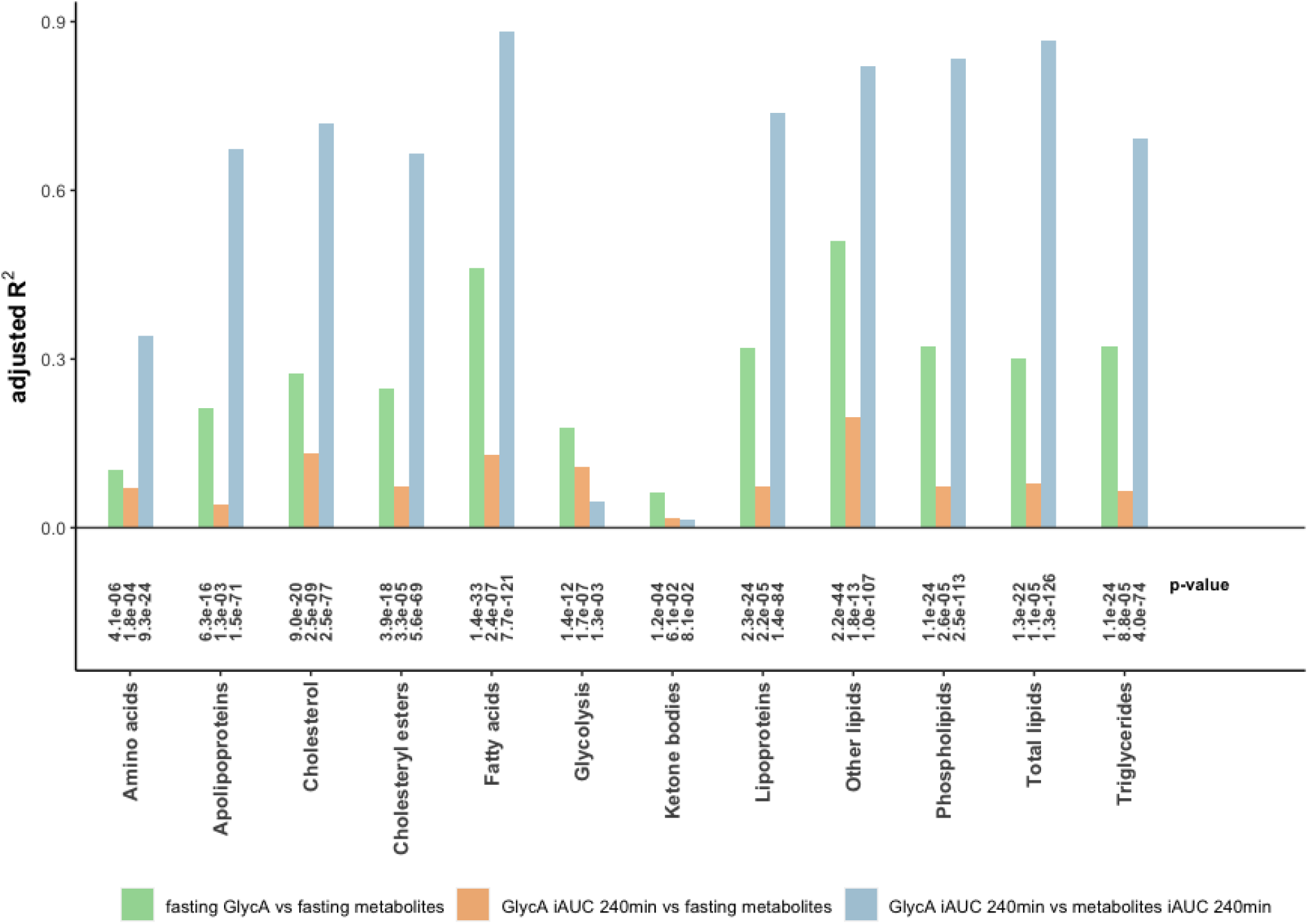
GlycA concentration versus different groups of metabolites at fasting and postprandial states. The results are from multivariable linear regression models, where the concentration of various groups of metabolites are included as predictors against fasting and postprandial levels of GlycA.

To assess the predictive potential of the fasting metabolome for postprandial inflammation, linear models were fitted between the fasting concentration of the same 12 groups of metabolites and GlycA_iAUC240min_ (Fig. 4). ‘*Other lipids*’ category was the most predictive, explaining 20% of the variance in postprandial inflammation, followed by ‘*cholesterol*’, ‘*glycolysis related metabolites*’, and ‘*fatty acids*’, each accounting for 11-13% of the variance. Subsequently, to assess the combined predictive utility of all the groups, Least Absolute Shrinkage and Selection Operator (LASSO) models were fitted between the fasting metabolites levels and GlycA_iAUC240min_. The model corresponded to only 14% of the variance in GlycA, with sphingomyelins, glucose, and alanine among the most important and stable features (Supplemental Fig. 3).

The interplay between the postprandial metabolome and postprandial inflammation was also evaluated, by fitting linear models between the iAUC_240min_ of the 12 groups of metabolites and GlycA_iAUC240min_ (Fig. 4). ‘*Fatty acids*’, several of lipid-related groups, ‘*cholesterol*’, and ‘*triglycerides*’ were all highly associated with postprandial inflammation, (*adj. R*^2^ : 0.66 - 0.88), whereas glycolysis-related-metabolites were less predictive (*adj. R*^2^ = 0.05 ; *P* = 1.3*e*^-3^).

### The modifying effect of anthropometric measures on the association between insulin resistance and GlycA

We examined how anthropometric measures modified the association between HOMA-IR and fasting GlycA levels. Propensity scores for insulin resistance were calculated using a combination of BMI and body fat percentage through linear models, and then dichotomized using median splits. Interaction analysis showed interactions between propensity score groups and GlycA levels on the inference towards HOMA-IR (*P* = 7.3*e*^-5^). The relation between fasting GlycA levels and insulin resistance (estimated by HOMA-IR) for high/low propensity groups are shown in Fig. 5. In the group with high propensity for insulin resistance, GlycA levels were significantly associated with HOMA-IR (*R* = 0.49 ; *P* = 2.5*e*^-9^). However, no association was observed in the low propensity group.

**Fig 5.**
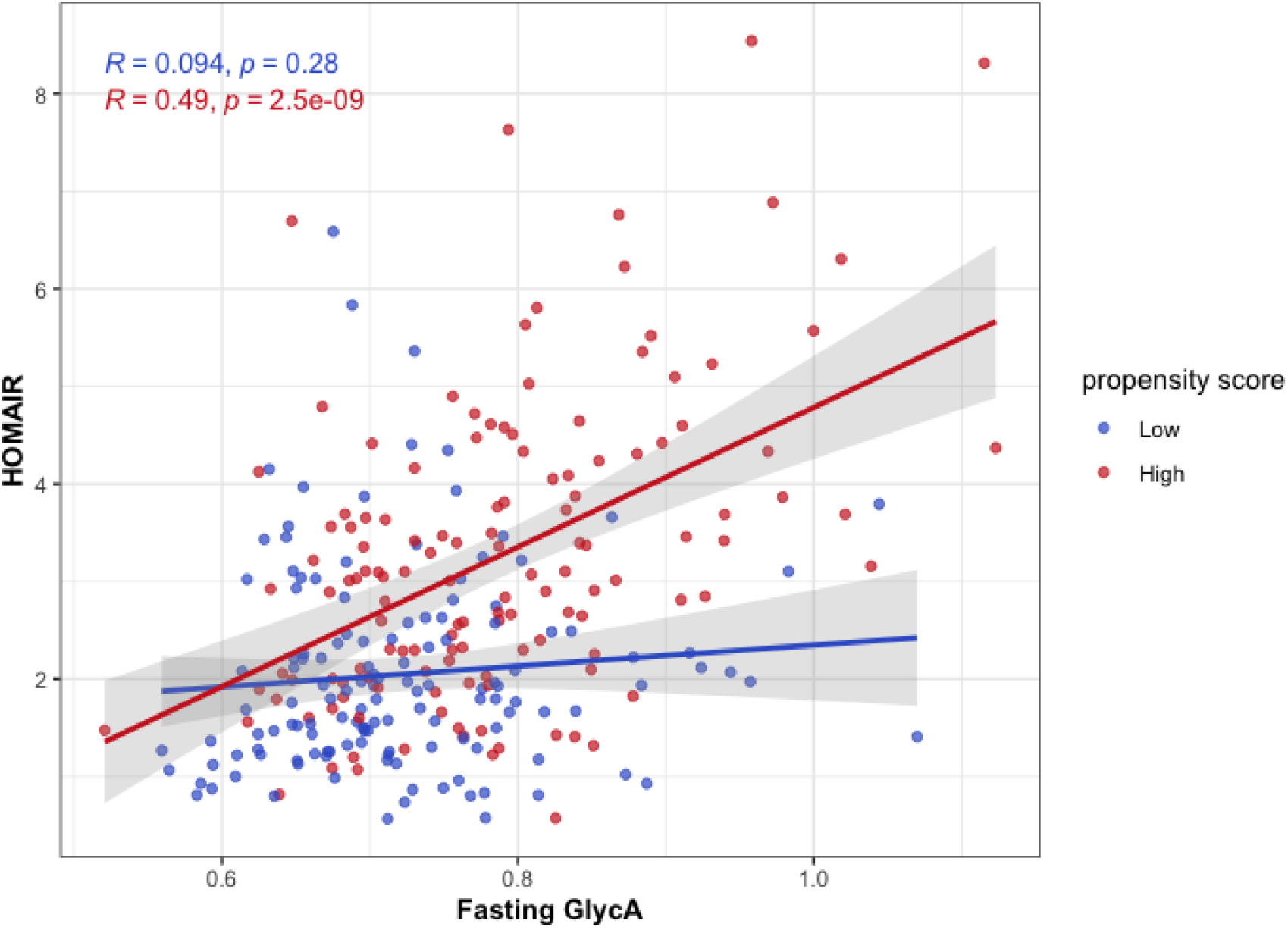
Association between HOMAIR and fasting GlycA levels, for groups with low and high propensity for developing insulin resistance. T propensity scores were calculated using BMI and body adiposity and dichotomized using the median value. The propensity score modifies t association between GlycA and HOMAIR ().

## Discussion

Our results showed differences between sexes in GlycA levels and postprandial dynamics. Females consistently showed higher GlycA concentrations at all time points, whereas males had more intense postprandial inflammatory responses. Most studies report greater postprandial lipemic and glycaemic response in males [26, 27], and given the known links between postprandial lipemia and glycemia with inflammation, our finding of greater postprandial inflammatory response in males was anticipated. During a mixed-nutrient meal challenge in the PREDICT (Personalised REsponses to DIetary Composition Trial) 1 study in a adult population, males not only showed greater postprandial inflammatory response, but also had significantly higher GlycA concentrations at fasting which is contrary to our observation [22]. Additionally, they report that GlycA concentrations are age-dependent, increasing with age. The disparity in age distribution between the two study populations (mean age = 45.6y) may account for the difference that we observed in our findings at the fasting state.

We observed that the fasting state chronic inflammation was more predictable by the investigated factors, than the transient postprandial inflammation. Fasting state chronic inflammation was associated with anthropometrics, blood pressure, insulin resistance, and life-style factors including alcohol consumption, smoking, physical activity, and stress, whereas transient postprandial inflammation showed fewer and weaker associations, with genetic risk, insulin resistance, blood pressure, and stress being the strongest determinants. Smoking and drinking did not show any associations with postprandial inflammation. However, both behaviours are known to impact postprandial lipemic response which can in turn induce inflammation [28]. It is possible that the duration of our study did not allow capturing the whole dynamics of postprandial lipemic-induced inflammation, as postprandial lipemic response peaks at 4-6 hours after ingestion [22, 29].

A number of the observed associations with fasting and postprandial inflammation were sex-specific. In males, waist-to-height ratio (proxy for visceral adiposity) was associated with a more intensified postprandial inflammatory response, highlighting the role of abdominal fat in less favourable postprandial response. Several of the interesting sex-specific associations were related to life-style factors. The association of heavier alcohol intake and smoking with increased fasting GlycA levels was much more significant in females. This indicates that females may be more susceptible to the adverse effects of alcohol and smoking. Another sex-specific association was related to blood pressure which is an established CVD risk factor [30, 31]. These associations were stronger in males in our cohort, aligning with many studies that report more favourable CVD risk factor profiles in women [32]. Although relations between GlycA and CVD have already been reported [19], identifying associations between systemic inflammation and subclinical phenotypes of CVD in our young generally healthy population, underscores the potentials of GlycA in preventive clinical research, offering insight into underlying pathological mechanisms decades before overt disease.

We observed that maternal smoking was associated with elevated fasting GlycA levels, even after adjusting for current smoking and social circumstances. Previous studies have demonstrated that foetal nicotine exposure can lead to long-term health impacts in the offspring and conditions such as T2D, obesity, and hypertension in the adulthood [33, 34]. Therefore, although we report the association between elevated fasting GlycA and pregnancy smoking for the first time, existing literature suggests evidence on the life-long detrimental effects of pregnancy smoking. Furthermore, we identify associations between fasting GlycA and other early life factors. Birth weight and being breastfed for at least 6 months of life corresponded to lower fasting GlycA levels. Similar observations have been reported for CRP, a conventional marker of inflammation [35]. Moreover, we observed that premature adiposity rebound was associated with higher fasting GlycA levels in males. Premature adiposity rebound has previously been associated with obesity in adulthood and worse cardiometabolic health later in life [36-38]. Based on previous evidence and our results, inflammation appears to be the endotypic mechanism linking early life factors to morbidities later in life.

BMI and body fat percentage are well known risk factors for insulin resistance. However, we hypothesized that the relation between inflammation and insulin resistance may depend on the level of risk and propensity to insulin resistance (estimated by BMI and body adiposity). We observed that propensity to insulin resistance had a modifying effect on the association between HOMA-IR and GlycA. Among individuals with high propensity to insulin resistance, fasting GlycA levels were significantly associated with HOMA-IR, whereas in the low propensity group, there was no such association. This indicates that in individuals with higher risk for developing insulin resistance due to overweight and obesity, systemic inflammation should be targeted to improve metabolic health. Specially, in cases where treating obesity itself is difficult due to various reasons, focusing on parallel strategies to lower systemic inflammation can be considered. Inflammation has been recognized as a contributing factor to poor metabolic health in individuals with obesity. Research has shown that those with obesity who exhibit lower systemic inflammation (assessed by other markers than GlycA) are shown to yet sustain their metabolic health, referred to as metabolically healthy obesity [39, 40].

We used machine learning (LASSO) to test the utility of the fasting metabolome for predicting postprandial inflammation. However, the fasting metabolome was not a strong predictor of the meal-induced inflammatory response. On the other hand, postprandial metabolome was highly predictive of postprandial inflammation. This suggests that the dynamic postprandial inflammation is regulated by the transient postprandial metabolome rather than the fasting. The association between lipaemic response and postprandial inflammation was much stronger compared to the glycaemic response, aligning with previous studies [21, 22]. This pattern was also present in the fasting state, with lipaemia-related metabolites accounting for nearly twice the variance of fasting inflammation compared to glycolysis-related metabolites.

One challenging association to interpret was related to vitamin D. We observed positive associations between postprandial inflammation and vitamin D levels, contrasting with the negative association at fasting. Although there is biological plausibility for vitamin D to modulate inflammation [41, 42], and vitamin D levels are generally reported to be inversely associated with inflammation, vitamin D is not an independent predictor of inflammation. This relationship is complex and influenced by multiple factors, making it difficult to isolate the effect of vitamin D alone [43, 44].

Our results suggest that initiatives aimed at encouraging breastfeeding and enhancing birth outcomes could yield significant clinical benefits by reducing chronic inflammation and lowering the risk of metabolic diseases in adulthood. Our results also highlight the importance of maintaining a healthy and active lifestyle, by avoiding harmful habits such as smoking and excessive alcohol consumption, being physically active, and managing stress. Smoking and excessive alcohol consumption seem to be increasing systemic inflammation more in females than males. For physical activity, we observed that it was only the duration of vigorous activity that was associated with attenuated fasting-state inflammation in our cohort. Another study has reported that only vigorous activity improved psychological well-being in adolescents [45]. These observations suggest a potential mechanistic link between overall well-being and systemic inflammation. Consequently, reducing systemic inflammation through vigorous physical activity or clinical practices can be considered to improve the overall well-being of adolescents.

The evidence is mounting from randomized control trials on the feasibility of reducing GlycA levels, for instance through lifestyle interventions and exercise [46-48]. The significant interindividual variability in fasting and postprandial inflammation, emphasizes the importance of adopting precision and personalized approaches for mitigating inflammation in preventive healthcare. To achieve this aim, research endeavours including ours, can aid in discerning factors that influence systemic inflammation at various metabolic stages, and guiding the development of tailored strategies to address inflammation in a more targeted and individualized manner.

We acknowledge the strengths and limitations of our study. Our study focused on a cohort of adolescents, whereas most other studies include mixed age groups or focus on older adults. Identifying the determinants of inflammation at the generally healthy adolescent stage can pave the road towards designing early and more efficient preventive strategies and life-style interventions. Furthermore, we utilized GlycA biomarker to evaluate inflammation, which is robust and consistently responsive to meal intake. This enabled us to accurately quantify postprandial inflammation. Regarding the limitations, the 4-hour duration of our study did not allow capturing the whole dynamics of postprandial lipemic-induced inflammation which peaks at 4-6 hours after ingestion. In addition, we did not find any associations between asthma and allergic rhinitis with either fasting or postprandial inflammation measured as GlycA, despite the known involvement of inflammation in the pathology of these conditions [1, 2, 49]. This could be caused by the mostly mild to moderate asthma cases in the cohort, which are all under close follow-up and well controlled. The other limitation of the study was the lack of control for smoking prior to the meal challenge. We observed that some of the daily smokers had considerably higher baseline GlycA levels that contrary to most of the population, decreased during the postprandial phase. We believe this was due to post-smoking inflammation [50, 51] interfering, where for individuals who had smoked in the hours immediately prior to taking the meal challenge, the smoking-induced inflammatory response hindered capturing the true level of postprandial inflammation. Therefore, we advise that smoking prior to taking samples for inflammation studies or undertaking meal challenges must be restricted.

In conclusion, this comprehensive study identified some of the determinants of fasting and postprandial inflammation from the lifespan, with a focus on pinpointing modifiable factors to mitigate inflammation. Our findings hold potential for advancing precision nutrition practices by enabling the phenotyping of individuals based on the identified determinants and subsequently their risk for elevated systemic inflammation. Future research should focus on leveraging our results to develop targeted dietary and lifestyle interventions aimed at reducing systemic inflammation. Moreover, investigating the causal relationships of the identified risk factors with inflammatory diseases can be of interest.

## Data Availability

All data produced in the present study are available upon reasonable request to the authors

## Acknowledgement

COPSAC is supported by both private and public research funds, all of which are detailed on our website, www.copsac.com. Core support has been provided by the Lundbeck Foundation, the Danish Ministry of Health, the Danish Council for Strategic Research, and the Capital Region Research Foundation. No pharmaceutical companies were involved in this study. The funding agencies had no role in the design or conduct of the study, nor in the collection, management, interpretation of the data, or in the preparation, review, or approval of the manuscript. Additionally, MAR received funding from the Novo Nordisk Foundation (NNF21OC0068517). Finally, we extend our sincere gratitude to Dr. Sarah Berry from King’s College London for her insightful comments and constructive feedback on the manuscript.

## Methods

### Study population

The participants were part of the COPSAC2000 cohort, which is a prospective clinical mother-child cohort study of 410 children of asthmatic mothers [24]. The participants were deeply-phenotyped with 17 scheduled visits to the COPSAC research clinic from birth till age 18. At the 18-year visit, 298 of the participants were given an Oral Glucose Protein Lipid Tolerance Test (OGPLTT).

### The meal challenge and blood sample collection

298 out of the 410 participants of the COPSAC2000 cohort underwent and completed a meal challenge at the 18-year visit. There were various reasons for the dropouts, including: vomiting, fear of needles, failing to insert IV, lactose intolerance, vegan diet, diabetes or other diseases, or plain refusal to participate in this part of the 18-year visit. The participants had fasted for at least 8 hours before the challenge. After insertion of an intravenous catheter and collecting baseline (fasting) blood samples, they received an Oral Glucose-Protein-Lipid Tolerance Test (OGPLTT). The meal was a hot beverage, consisting of 75 g glucose, 60 g palm oil, and 20 g skimmed milk powder, adding up to a total caloric content of ∼947 kcal. This corresponded to ∼33% of daily caloric expenditure of the participants (∼ 3000 kcal), adjusted for gender and height. Pure vanilla powder was added to improve the taste, and 1.5 g paracetamol was added to assess ventricular passage time. The meal was consumed within a maximum of 30 min, and preferably 15 min. Once half of the meal was consumed, the timer was set for blood draws at 15, 30, 60, 90, 120, 150 and 240 min postprandially.

### NMR biomarker profiling

Circulating levels of Glycoprotein acetylation (GlycA) was measured for fasting and postprandial blood samples as part of a broader metabolic profiling, using a targeted high-throughput NMR metabolomics platform (Nightingale Health Ltd., Helsinki, Finland).

### Extracting metabolic features

In this study, GlycA levels measured from the baseline fasting blood samples were used as fasting-state GlycA levels. To extract postprandial features, incremental area under the curve (iAUC) of GlycA profiles were calculated as defined previously [52], using mainly basic R commands and ‘ggplot2’ package. First, smoothed curves were fitted to individual profiles, and then after subtracting the fasting (baseline) GlycA levels from the fitted curve, the area under the curve was calculated as the summation of positive increments.

### Data analysis

To investigate the association of risk factors with GlycA levels at fasting and postprandially, (multiple) linear regression models were fitted between risk factors and GlycA levels. When deemed appropriate, adjustment variables were included in certain models. Moreover, in cases where stratification by sex was warranted due to biological factors (e.g. Tanner score), or where stratification would reveal additional sex-specific differences compared to non-stratified models, results from the stratified models were reported.

Effect sizes were estimated using (adjusted) coefficient of determination (*R*^2^) for univariable and multiple linear models. On models that included adjustment variables, we subsequently performed an Analysis of Variance (ANOVA), to calculate ‘eta squared’ (males showed positive association *η*^2^: sum of squares of the predictor/total sum of squares) as an estimate of the effect size. Due to the descriptive nature of this study, we report inference as uncorrected *p-values*.

The association of risk factors with postprandial GlycA was investigated at two time-points: 90 and 240 min. 90 min was chosen based on the median GlycA concentration profiles, for the maximum postprandial inflammation to have reached in most participants, and 240 min was chosen as it is the last monitored time point and metabolites such as triglycerides that can affect GlycA levels will have their highest concentration.

For anthropometrics and genetic risk (polygenic risk scores) backward selection was used to identify the most significant and relevant predictors to include in the regression models, by iteratively removing less important variables. The composition of all the models and detailed results are presented in Supplemental Table 2.

To explore the relation between fasting concentrations of 12 groups of key metabolites and fasting and postprandial inflammation, multivariable linear regression was used with metabolites fasting concentrations as the predictors and GlycA levels as the response variables, separately for fasting GlycA and GlycA_iAUC240min_. Similar models were run using metabolites postprandial concentrations (iAUC_24min_) as predictors and GlycA_iAUC240min_ as the response, to investigate the interplay between the postprandial metabolome and postprandial inflammation. Subsequently, to assess the utility of all the 12 groups (fasting metabolome) for predicting postprandial inflammation, Least Absolute Shrinkage and Selection Operator (LASSO) models were fitted between the fasting metabolites levels and GlycA_iAUC240min_, using ‘Caret’ package in R.

To investigate how anthropometric measures modified the association between HOMA-IR and fasting GlycA levels, propensity scores for insulin resistance were calculated using a combination of BMI and body fat percentage, through linear models. The propensity score was then dichotomized using the median value. Subsequently, we did interaction analysis between propensity score groups and GlycA levels using linear models and ANOVA. The relation between fasting GlycA levels and HOMA-IR was shown by fitting linear models, for the high and low propensity groups.

### Phenotyping and characterizing (risk) factors

#### Gestation and Birth

Information regarding alcohol consumption and smoking habits during pregnancy was gathered through questionnaires. For the purposes of this study, smoking and alcohol consumption were characterized as engaging in either practice in any of the trimesters, as dichotomous variables. Data on antibiotics intake during pregnancy and preeclampsia was obtained from the Danish National Health Registry.

Data related to birth, including gestational age, birth weight, and Apgar scores were obtained from the Danish National Health Registry. Moreover, cord blood was collected after birth and frozen. To obtain the cytokines composition of the samples, samples were shipped on dry ice to be analysed at the Department of Clinical and Experimental Medicine, Linköping University, Sweden. After thawing, the levels of IP10, I-TAC, TARC, and MDC were measured, using an in-house multiplexed Luminex assay. The limit of detection was 6 pg/ml for IP10, 28 pg/ml for I-TAC, and 2 pg/ml for TARC and MDC. All samples were analysed in duplicates, and the sample was re-analysed if the coefficient of variation (CV) was >15% [53].

#### Early Life

##### Breastfeeding

Breastfeeding information was acquired through interviews with the families. For the purpose of this study, breast feeding was defined as a dichotomous variable, indicating whether infants were breastfed for a minimum of the initial six months of life. This classification encompassed cases of exclusive breastfeeding, as well as instances where breastfeeding occurred in conjunction with other nutritional sources.

##### Breast milk fatty acids

Breast milk samples ranging from 2 to 5 mL were collected from mothers approximately one month postpartum. The timing of sample collection during feeding was at the mothers’ discretion, as the fatty acid composition remains consistent throughout feeding. Aliquots of 2 mL of milk were treated with 0.01% 2,6-di-tert-butyl-4-methylphenol (Sigma Chemical, St. Louis, MO) and stored at −80°C. All samples were analysed within one year of collection. Lipids were extracted from 1 mL of milk and subsequently methylated using potassium hydroxide in methanol. The resultant fatty acid methyl esters were extracted with heptane and separated by gas–liquid chromatography (Hewlett-Packard, Waldbronn, Germany), following established protocols. Peaks, ranging from lauric acid (12:0) to DHA, were identified based on the retention times of commercial standards (Nu-Chek Prep, Elysian, MN). Ninety-seven percent of the fatty acids within this range were successfully identified [54]. The content of individual fatty acids or fatty acid classes was expressed as a percentage by weight of the total fatty acid content. The list of the measured fatty acids is included in Supplemental Tables 6.

##### Breast milk cytokines

Breast milk samples, approximately 10 mL each, were collected from participants one month postpartum at the COPSAC research facility. The samples were aliquoted and stored at -80°C pending analysis. Upon thawing, the fatty layer and cellular components of the breast milk were removed via centrifugation at 680g for 10 minutes at 4°C. Subsequently, the samples were further centrifuged at 10,000g for 30 minutes at 4°C, resulting in a translucent liquid which was then aliquoted and stored at -80°C until analysis. The analysis focused on detecting 19 proinflammatory and immunoregulatory cytokines and chemokines, which are important in the context of eczema and asthma immunopathology. Custom-made ultrasensitive multiplex assays from Meso Scale Discovery (Gaithersburg, MD, USA) were employed for the analyses. The samples were analysed in duplicates in a randomized sequence and read using a Sector Imager 6000 (MSD). For each immune mediator, zero values were substituted with half the lowest detectable level. The calibration levels for the chemokine assays ranged from 20,000 to 1.22 pg/mL, with no observed interference at these concentrations [55]. The list of the targeted cytokines and chemokines can be found in Supplemental Table 7.

##### Infection burden

Infection burden in the first three years of life was estimated at the scheduled 6-monthly visits from birth until 3 years of age, as well as additional acute care visits where our paediatricians assessed the child and interviewed the parents about any illnesses, symptoms, duration, medication, and vaccinations since the last visit. When necessary, the physician added additional clinical information. In case of missed visits, the parents were interviewed at the subsequent visit on infectious episodes. Infections were classified according to the International Classification of Diseases, 10th Revision22 and stored in a designated database. The infections were classified in the following groups: 1-Upper respiratory tract infections (URTIs): common cold, tonsillitis, pharyngitis, otitis media, and croup; 2-Lower respiratory tract infections (LRTIs): pneumonia and bronchiolitis; 3-Gastrointestinal infections (GIs): GI, diarrhoea, and vomiting; and 4-Isolated fever and other infections. For the current study, the overall incidence of infections (the 4 groups combined) were used as an estimate of infection burden during the first 3 years of life.

##### Blood cytokines at 7 years

Cytokine levels, including C-Reactive Protein (CRP), Interleukin-1 beta (IL-1β), Interleukin-6 (IL-6), Interleukin-8 (IL-8), and Tumour Necrosis Factor-alpha (TNF-α), were measured in blood samples collected from participants at age 7. Venous blood samples were drawn, centrifuged to separate plasma from blood cells, and immediately stored at -80°C until analysis. Samples were transported on dry ice to the laboratory, where inflammatory biomarkers were quantified using high-sensitivity ELISA assays based on electro-chemiluminescence. A 4-plex setting was employed for IL-1β, IL-6, CXCL8, and TNF-α, while hs-CRP was measured using a single high-sensitivity assay from Meso Scale Discovery. The lower limit of detection (LLOD) for the assays was 0.005 ng/mL. Each sample was analysed in duplicates using the Sector Imager 6000 (Meso Scale Discovery®, Gaithersburg, MD, USA). The detection limits (mean signal from blanks +3SD) were 9.54 pg/mL for hs-CRP, 0.15 pg/mL for IL-1β, 0.17 pg/mL for IL-6, 0.09 pg/mL for CXCL8, and 0.08 pg/mL for TNF-α.

##### Growth trajectories

Growth trajectories were modelled using latent class growth trajectories from z-BMI scores, modelled longitudinally from children at 1 week to 18 years. We considered linear latent class models between 2-5 and selected the model with 4 classes based on model fit criteria (AIC and BIC) and biological considerations.

##### Adiposity rebound

Adiposity Rebound is identified by a shift in the ratio of the velocities of log(weight) to log(height) from less than 2 to 2 or more. To investigate the “age at AR”, children with at least three recorded measurements of weight and height around the age of 5 were considered.

##### Tanner score

Tanner staging classification system was used to assess the development of the participants into puberty at 12 years, based on the development of external genitalia and secondary sex characteristics, including phallus, scrotum, and testes volume in males, breasts in females, and pubic hair in both males and females [56].

##### Siblings

Data regarding the presence of older siblings was gathered when the participants reached 7 years of age.

#### Current

##### Anthropometrics

Anthropometrics were assessed at each clinical visit. Waist circumference was measured using a tape, placing it around the navel as a reference point and averaging two measurements taken during inspiration and expiration. Height was gauged using a stadiometer, calibrated annually, and weight was recorded without clothes on calibrated digital scales. Body composition analysis at age 18 involved Bioelectrical Impedance Analysis (BIA) using a Tanita scale (Health monitor, version 3.2.7). This provided measurements of muscle mass (kg), fat mass (kg), and bone mass (kg). Derived metrics included fat-free mass (FFM) (body weight minus fat mass), fat mass index (fat mass divided by height squared), and fat-free mass index (fat-free mass divided by height squared in meters).

##### Polygenic risk scores

The construction of Polygenic Risk Scores (PRS) was facilitated using the software package PRS-CS (Polygenic Risk Score Continuous Shrinkage), which regularizes the effects of single nucleotide polymorphisms (SNPs) using a shrinkage prior. PRS-CS employs a linkage disequilibrium (LD) reference panel derived from European ancestry samples in the 1000 Genomes Project. The polygenicity parameter (phi) was automatically estimated within this framework. Following the adjustment of SNP effects using PRS-CS, PLINK2 software was utilized to consolidate SNP effects into individual PRSs. These scores were then standardized to have a mean of 0 and a standard deviation of 1 for each phenotype. A shortlist of 15 PRS (associated with T2D, metabolic disorder, inflammation, CVD, and inflammatory disease) were selected to be analysed against fasting and postprandial inflammation. The complete list of these PRS is presented in Supplemental Table 4.

##### Smoking and alcohol habits

Smoking habits and alcohol consumption were documented at the 18 years visit using a structured interview. For our analysis, we classified individuals who reported daily or weekly smoking as ‘smokers’, and weekly alcohol consumption exceeding 10 units as heavy drinking habits.

##### Accelerometer

Activity and sleep data were obtained at 18 years, from accelerometer readings (Actigraph GT3X+, 30Hz) over a 14-day period, analysed using the GGIR package in R (version 2.9.0). To ensure accuracy, the autocalibration feature in the GGIR package was used for calibrating individual accelerometers. For quality control, 7 participants were discarded due to auto-calibration issues. Analysis excluded participant data with less than four days of recordings, and days with less than 16 hours of wear time were removed from individual datasets. The analysis involved calculating the mean daily vector magnitude (mg) over 5-second epochs. This data was used to quantify sedentary time (activity below 30 milli-G’s (mg)), light activity (30-100mg), moderate activity (100-400mg), and vigorous activity (over 400mg). Sleep onset was determined as the average time sleep began, and sleep duration was calculated as the average length of sleep in hours.

##### Stress

Level of stress experienced by the participants was assessed using the Depression, Anxiety and Stress Scale (DASS-21) Questionnaire, at 18-year visit.

##### Social circumstances

Social circumstances of the participants was estimated using the first principal component of household income, maternal education level, and maternal age at birth.

#### Paraclinical measures and Clinical endpoints

##### Insulin resistance (HOMA-IR)

Insulin sensitivity was estimated at 18 years, based on the fasting insulin and glucose levels in blood, using the homeostatic model assessment [57].

##### Blood Pressure

Systolic and diastolic blood pressure were measured by research assistants during the 18-year clinical visit. Three measures were taken when the participants were seated and relaxed, and the average of the second and third measurements was used in this analysis.

##### Blood biochemistry measures

Blood samples were collected during the 18-year visit. These samples were taken at fasting state and were analysed the same day, for an extensive list of clinical biochemistry measures. The list of the included tests can be found in Supplemental Table 3.

##### Asthma

Asthma was diagnosed based on predefined internationally recognized guidelines [58, 59], including 4 mandatory criteria: 1) Diary-verified recurrent troublesome lung symptoms (referred to as “recurrent wheeze”), defined as 5 episodes of minimum 3 consecutive days within 6 months or 4 consecutive weeks with symptoms; 2) Symptoms judged by the COPSAC paediatricians to be typical of asthma (e.g. exercise induced symptoms, prolonged nocturnal cough, recurrent cough outside common cold, symptoms causing waking at night); 3) Intermittent need of rescue inhaled β2-agonist; and 4) Response to a 3-month trial of inhaled corticosteroids initiated when the criteria for recurrent troublesome lung symptoms were met and relapse after cessation.

##### Allergic rhinitis

Allergic rhinitis was diagnosed based on parental interviews (not questionnaires) on history of symptoms performed at the COPSAC research unit. Allergic rhinitis was defined as bothersome and recurrent sneezing, blocked, itchy or runny nose severely affecting the wellbeing of the child in the past 12 months in periods without accompanying common cold or flu, and congruence between symptoms, relevant exposure, and positive SPT and/or sIgE [60].

## Ethical approval

The study was approved by the Ethics Committee in Region Hovedstadens in Denmark (H-16040846) and the Danish Data Protection Agency (2015-41-3696).

## Notes

### Competing Interest Statement

The authors have declared no competing interest.

