## Supplementary material for "Systemic inflammation during fasting and postprandial states: a comprehensive study of key determinants in a deeply characterized cohort of young adults": Supplementary_material.docx

Supplemental Table 1. a) Descriptive characteristics of COPSAC2000 cohort, and the participants versus non-participants of the meal challenge, b) sex-stratified characteristics of the meal challenge participants.

**Baseline characteristics**

| **Characteristic** | **N** | **Overall, N = 410**^1^ | **non-participated, N = 112**^1^ | **participated, N = 298**^1^ | **p-value**^2^ |
| --- | --- | --- | --- | --- | --- |
| **BMI** | 365 | 22.3 (20.0, 24.9) | 22.7 (20.7, 25.3) | 22.2 (19.9, 24.6) | 0.12 |
| **Weight** | 365 | 68 (61, 77) | 69 (62, 77) | 67 (61, 78) | 0.3 |
| **Height** | 365 | 1.75 (1.68, 1.81) | 1.75 (1.68, 1.82) | 1.75 (1.68, 1.81) | >0.9 |
| **Body fat percentage** | 348 | 23 (17, 29) | 23 (17, 29) | 23 (17, 29) | 0.7 |
| **Muscle-to-fat ratio** | 348 | 3.13 (2.31, 4.75) | 3.20 (2.29, 4.76) | 3.12 (2.31, 4.71) | 0.7 |
| **Fat mass index** | 348 | 4.99 (3.53, 7.00) | 5.01 (3.50, 7.33) | 4.95 (3.59, 6.87) | 0.9 |
| **HOMA-IR** | 329 | 2.33 (1.60, 3.46) | 2.73 (2.12, 6.39) | 2.27 (1.54, 3.40) | <0.001 |
| **Systolic blood pressure** | 363 | 115 (109, 122) | 115 (110, 120) | 115 (109, 123) | >0.9 |
| **Diastolic blood pressure** | 363 | 71.0 (68.0, 75.5) | 71.5 (68.5, 75.5) | 71.0 (67.5, 75.1) | 0.6 |
| *^1^* Median (IQR) | | | | | |
| *^2^* Wilcoxon rank sum test | | | | | |

| Participants characteristics stratified by sex | | |
| --- | --- | --- |
|  | **Female** | **Male** |
| Number of participants | 155 | 144 |
| Weight (kg) | 65.23 (12.34) | 75.34 (13.88) |
| BMI (kg/m2) | 22.98 (4.32) | 22.79 (3.91) |
| Height (cm) | 1.69 (0.06) | 1.82 (0.07) |
| Waist (cm) | 79.13 (11.44) | 81.30 (11.38) |
| Hip circumference (cm) | 90.22 (10.67) | 87.09 (10.87) |
| Waist to hip ratio | 0.88 (0.09) | 0.94 (0.08) |
| Muscle mass (kg) | 43.11 (5.39) | 57.79 (7.10) |
| Body fat percentage | 29.07 (6.23) | 18.19 (6.09) |
| Muscle to fat ratio | 2.45 (0.68) | 4.76 (1.60) |
| Blood glucose (mg/dL) | 96.92 (7.86) | 97.97 (6.76) |
| Insulin (mU/L) | 12.19 (6.12) | 9.72 (5.10) |
| HOMA-IR | 2.92 (1.56) | 2.33 (1.21) |

Supplemental Table 2. list of the risk factors, the input and adjustment variables for each model, and the linear model results

*See ‘Supplemental_Table2.xlsx’*

Supplemental Table 3. List of blood biochemistry tests.

*See ‘Supplemental_Table3.xlsx’*

Supplemental Table 4. List of investigated polygenic risk scores (PRS) versus fasting and postprandial metabolome

*See ‘Supplemental_Table4.xlsx’*

Supplemental Table 5. The linear models of fasting and postprandial GlycA levels versus fasting and postprandial metabolome

*See ‘Supplemental_Table5.xlsx’*

Supplemental Table 6. List of the measured breast milk fatty acids at approximately one month postpartum.

| **Fatty acids** | **Abbreviations** |
| --- | --- |
| Caprylic Acid (SFA) | C12:0 |
| Myristic Acid (SFA) | C14:0 |
| Pentadecanoic Acid (SFA) | C15:0 |
| Palmitic Acid (SFA) | C16:0 |
| Margaric Acid (SFA) | C17:0 |
| Stearic Acid (SFA) | C18:0 |
| Arachidic Acid (SFA) | C20:0 |
| Total Saturated Fatty Acids (SFA_TOT) | SFA_TOT |
| Palmitoleic Acid (MUFA) | C14:1-n5 |
| Palmitelaidic Acid (MUFA) (Possibly) | C16:1-n7 |
| Heptadecenoic Acid (MUFA) (? Specific Isomer) | C17:1? |
| Oleic Acid (MUFA) | C18:1-n9 |
| Vaccenic Acid (MUFA) | C18:1-n7 |
| Gadoleic Acid (MUFA) | C20:1-n9 |
| Erucic Acid (MUFA) | C22:1-n9 |
| Total Monounsaturated Fatty Acids (MUFA_TOT) | MUFA_TOT |
| Linoleic Acid (PUFA) | C18:2-n6 |
| Gamma-Linolenic Acid (PUFA) | C18:3-n6 |
| Eicosadienoic Acid (PUFA) | C20:2-n6 |
| Eicosatrienoic Acid (PUFA) | C20:3-n6 |
| Arachidonic Acid (PUFA) | C20:4-n6 |
| Adrenic Acid (PUFA) | C22:4-n6 |
| Docosapentaenoic Acid (PUFA) | C22:5-n6 |
| Total n-6 Polyunsaturated Fatty Acids (n6_TOT) | n6_TOT |
| Alpha-Linolenic Acid (PUFA) | C18:3-n3 |
| Eicosapentaenoic Acid (PUFA) | C20:5-n3 |
| Docosapentaenoic Acid (n-3) (PUFA) | C22:5-n3 |
| Docosahexaenoic Acid (PUFA) | C22:6-n3 |
| Total n-3 Polyunsaturated Fatty Acids (n3_TOT) | n3_TOT |
| Total Polyunsaturated Fatty Acids (PUFA(y+ad)) | PUFA(y+ad) |
| Ratio of n-6 to n-3 Fatty Acids (n6/n3) | n6/n3 |
| Linoleic Acid to Alpha-Linolenic Acid Ratio (LA_ALA) | LA_ALA |
| Long-Chain Polyunsaturated Fatty Acids (PUFA_LCP) | PUFA_LCP |
| Trans Fats (TRANS) | TRANS |
| Unidentified Fatty Acids (UIDENT) | UIDENT |
|  | C12:0 |

Supplemental Table 7. List of the measured breast milk cytokines and chemokines at approximately one month postpartum.

**Cytokines and chemokines**

Eotaxin-3

GRO-a (Growth-regulated oncogene alpha)

IL-8 (Interleukin-8)

IP-10 (Interferon-gamma-induced protein 10)

MCP-1 (Monocyte Chemoattractant Protein-1)

MDC (Macrophage-Derived Chemokine)

MIP-1β (Macrophage Inflammatory Protein-1 beta)

RANTES (Regulated on Activation, Normal T Cell Expressed and Secreted)

TARC (Thymus and Activation-Regulated Chemokine)

TSLP (Thymic Stromal Lymphopoietin)

IFN-γ (Interferon-gamma)

IL-10 (Interleukin-10)

IL-13 (Interleukin-13)

IL-17 (Interleukin-17)

IL-1β (Interleukin-1 beta)

IL-4 (Interleukin-4)

IL-5 (Interleukin-5)

TNF-α (Tumor Necrosis Factor-alpha)

TGF-b1 (Transforming Growth Factor beta-1)

Supplemental Figure 1.
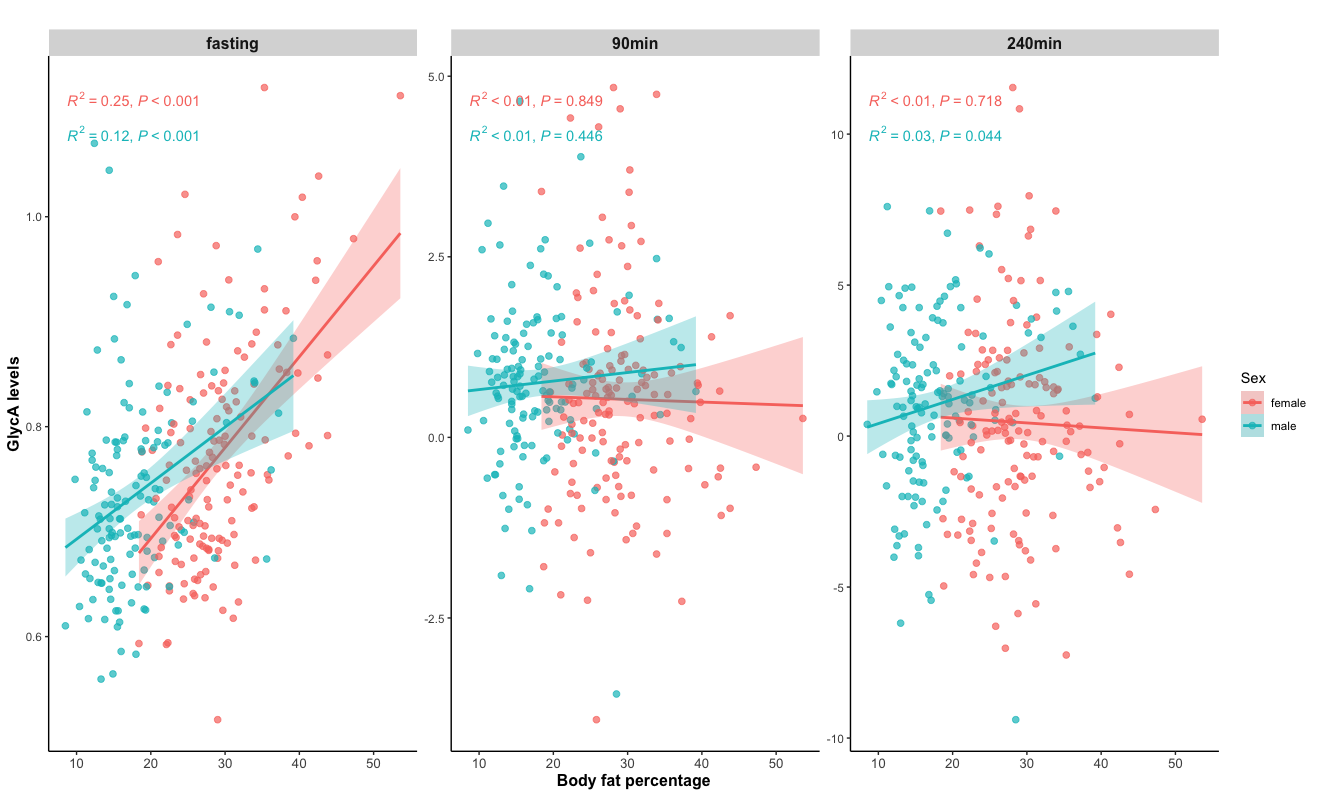
 Body fat percentage versus fasting and postprandial GlycA levels.

Supplemental Figure 2. Determinants of postprandial GlycA at 90 min (iAUC90min). ‘Blue’ bars represent positive regression coefficients as acquired from univariable linear regression, while ‘Red’ bars represent negative regression coefficients; these correspond to positive and negative associations between the risk factor and GlycA levels, respectively. For risk factors with multivariable models, the bars are colored ‘Gray’, with no immediate direction of association obtained. The labeled components in the figure correspond to: A) gestation, B) birth, C) early-life, D) current, and E) clinical endpoints.

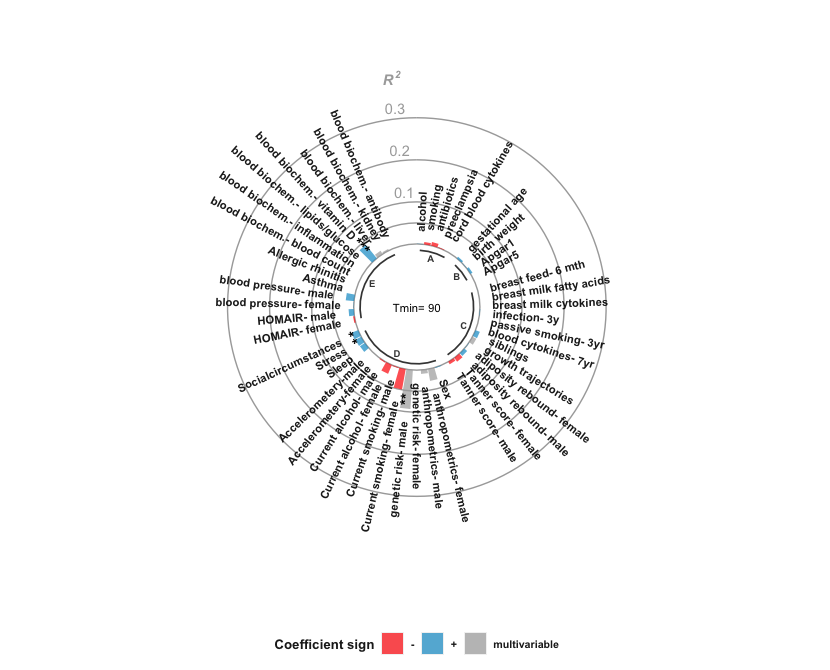

Supplemental Figure 3. LASSO models of baseline metabolites versus postprandial GlycA iAUC_240min_.

| 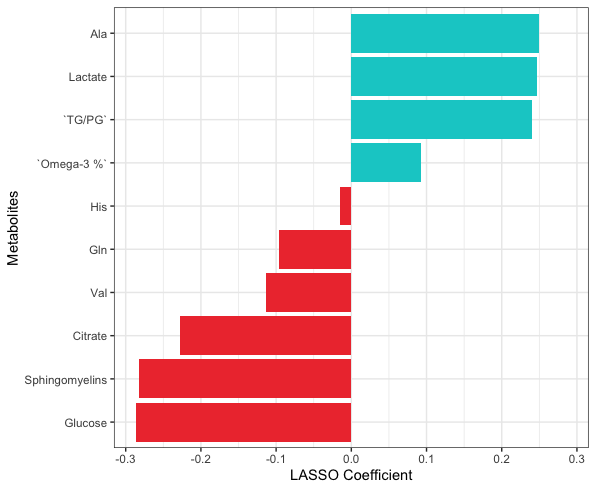 | 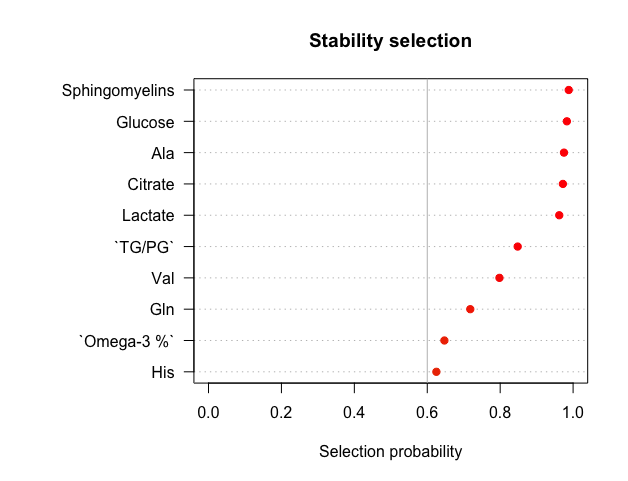 |
| --- | --- |
| 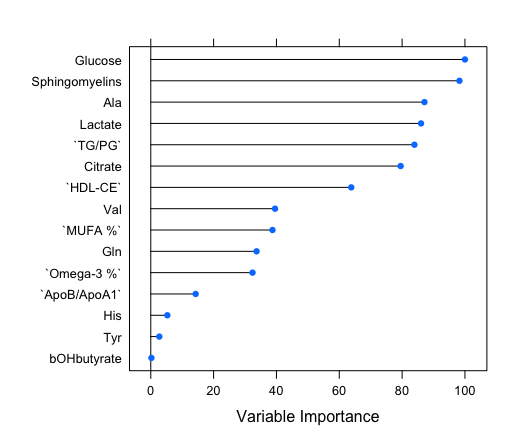 | Lambda \| R^2^ \| Root-mean-square error (RMSE) \| Mean absolute error (MAE); 0.069 \| 0.14$\pm$0.04 \| 1.91$\pm$0.17 \| 1.41$\pm$0.08 |
